# Estimating prevalence of immunocompromising conditions in Canada in 2022 using an administrative hospital database

**DOI:** 10.1101/2025.02.05.25321738

**Authors:** Philippe Finès

**Affiliations:** Formerly at Public Health Agency of Canada

## Abstract

**Background:** Immunocompromising conditions (ICs) are conditions that may be present at different moments in a person’s life and affect their health. In individuals with ICs, the body’s ability to effectively fight infections or mount a robust response to vaccination is weakened. It is thus important to understand the burden of ICs in part to inform public health policies on the procurement of vaccines for this population. However, no estimates of prevalence rates of ICs exist in Canada.

**Objective:** Our objective was to address this gap.

**Intervention:** We defined our list of ICs using different published sources and expert opinion. Using diagnoses coded with ICD-10 over a period of 14 years in the Discharge Abstract Database, we were able to estimate prevalence rates of ICs.

**Outcome:** For all ICs combined and conservatively assuming a hospitalization rate of 0.1, general population prevalence rates is estimated at 1.8% for females and 1.9% for males. Inflammatory bowel disease, severe liver disease and severe cancers are among the most frequent ICs at all age groups and for both genders. Functional asplenia and Primary Immunodeficient conditions are mostly present at young ages. Transplantation and renal dialysis are also among the most frequent ICs.

**Conclusion:** The prevalence of ICs estimated using DAD aligns with estimates obtained using other methods from other countries (2.7–6.6%) under certain assumptions for hospitalization rates. This work serves as the foundation for future investigation into obtaining more precise estimates of prevalence rates of ICs in Canada.

## I Introduction

### 1. Issue identification

Individuals with immunocompromising conditions (ICs) may have an increased risk of morbidity and mortality if under-immunized and may get serious adverse events if they receive inappropriate use of live vaccines^1,2^. Therefore, it is important to understand the burden of ICs to inform public health policies on the procurement and use of vaccines for this population^3,4^. ICs are characterized by a weakened state of the body’s ability to fight infections or mount a robust response to vaccination and may manifest as a result of a combination of congenital conditions, illnesses, and medications that suppress immune function. ICs present at birth are generally referred to as primary immunodeficient conditions (PID), whereas those that happen later in life are referred to as secondary immunodeficient conditions (SID); however, the distinction may be difficult to unravel^5^. Some medical conditions may not be inherently immunocompromising but may be considered as such when paired with immunosuppressive treatments. Conversely, treatments may mitigate against immunocompromising consequences of a medical conditions, as in the case of diabetes and HIV; only individuals with uncontrolled disease (i.e., not on treatment) are considered to be immunocompromised.

### 2. What is known to date

An estimate of the prevalence of ICs in the Canadian population does not currently exist. There are very few estimates of the burden of ICs in countries comparable to Canada: prevalence of ICs is 3.2% of the adult population in the United States (US), and 3.5% of the adult population in the United Kingdom (UK)^6^. It increased in the US from 2.7% in 2013 to 6.6% in 2021, according to Natjonal Health Interview Survey (NHIS) performed eight years apart^7,8^ In general, the prevalence of ICs appears to be increasing as screening for these conditions earlier in life has raised and life expectancy is growing.

### 3. Objective and rationale of the study

The purpose of this article is to address the gap regarding prevalence of ICs in Canada by analyzing administrative data.

## II Intervention

### 1. Setting/participants/interventions

A preliminary list of ICs was drawn from Varghese et al.^6^ and was used as a starting point to determine the burden of ICs in Canada. Medical advisors with the Public Health Agency of Canada (PHAC) were consulted to refine the list. The list was improved further, to include cancers identified as the most frequent^9^ and those defined as potentially immunocompromising by the Canadian Cancer Society^10^. The final version of the list of ICs is presented in Table 1 “List of ICs analyzed”. In this table, PIDs are regrouped into one category and SIDs are classified into two groups: Diseases and Treatments.

**Table 1.**
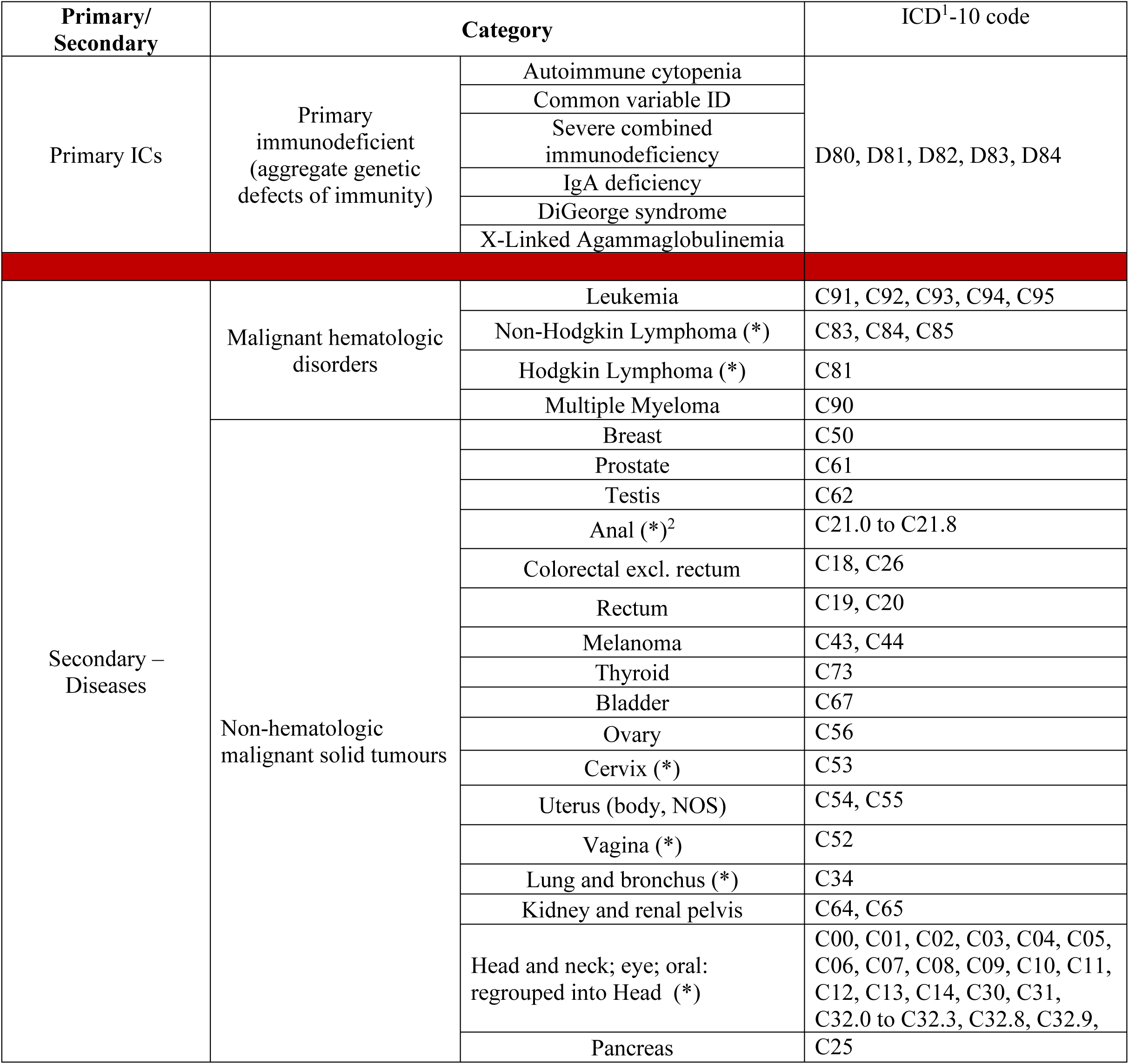
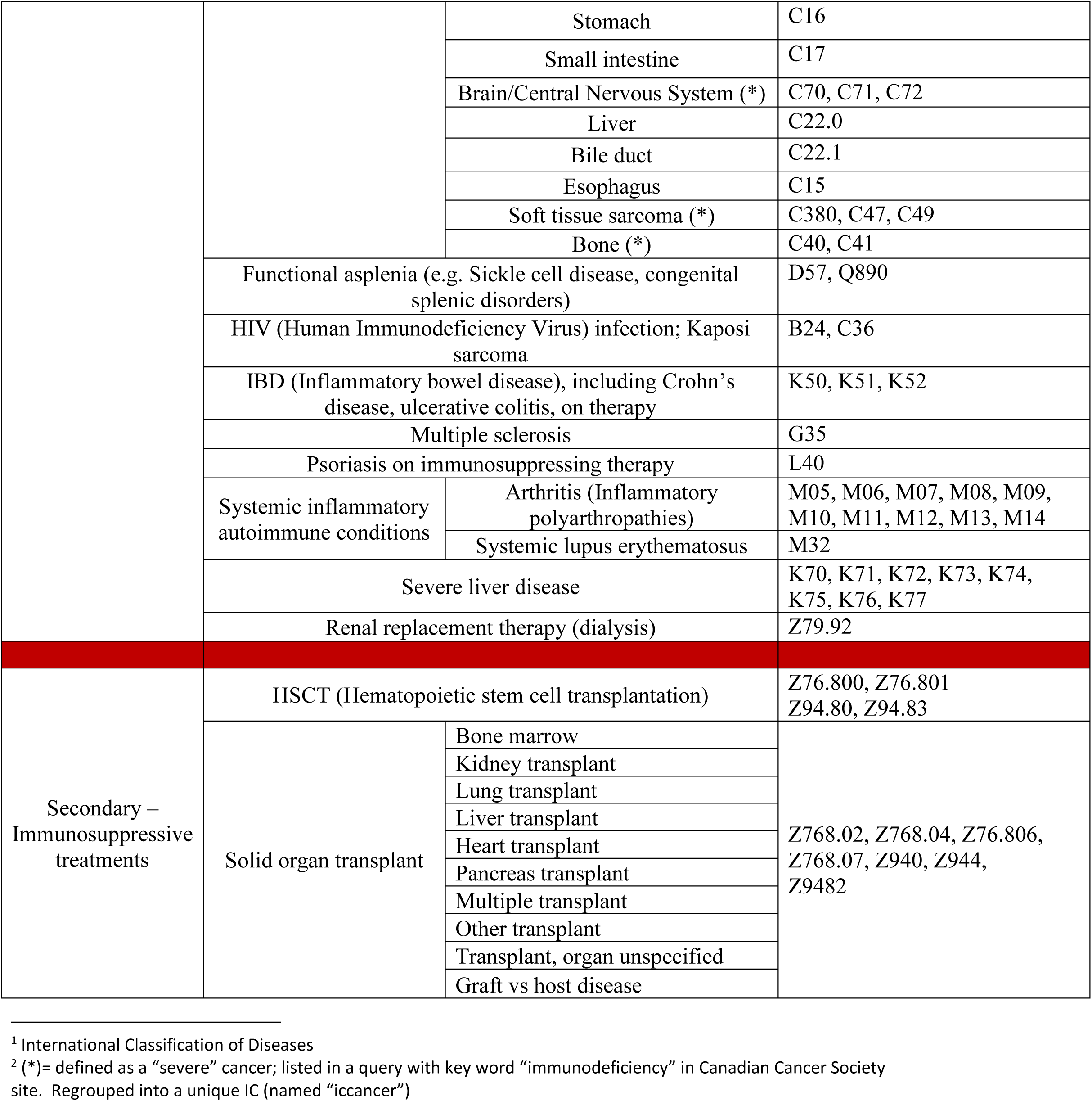
List of ICs analyzed.

Since no health surveys contained data on all the ICs we wanted to analyze, we used the Discharge Abstract Database (DAD)^11^, that contains all hospital visits in Canada since 2005 except for Québec. The diagnoses in DAD are coded by ICD-10^12^. Validation has already been performed on DAD data (sometimes with combination with other data sources), for example for the elaboration of an optimal algorithm for defining a disease^13-16^. In addition, four studies were restricted to Ontario as they combined DAD with databases specific to that province^17-20^.

We used DAD data to analyze the data of individuals who were hospitalized with at least one diagnosis of IC. For this study, we assumed that these data capture only the ICs severe enough to lead to hospitalization: ICs that did not lead to hospitalization were not captured in DAD.

The right-hand part of Table 1 “List of ICs” contains the diagnosis numbers (with ICD-10) associated to the list of ICs. It was not possible in DAD to convey all the nuances that would be required to define ICs. “Severe” cancers were defined as those that were related to immunodeficiency. Also, although immunosuppressive medications satisfy the definition of an IC, they were not included because no data in DAD contained the corresponding ICD-10 diagnosis for the years that were analyzed.

The DAD data contain 25 diagnoses per visit. For each IC, we considered a visit as being an occurrence of this IC if the corresponding diagnosis appeared as one of the 25 diagnoses of the visit. DAD data were read and transformed with the SAS software (Enterprise Guide, version 7.15HF3). Results were produced in SAS and completed with Excel: the final product was an Excel sheet.

### 2. Ethics review

Ethics review was not required as this study involved the analysis of aggregated, anonymized administrative data publicly available in Statistics Canada’s Research Data Centres and did not involve contacting individuals.

### 3. Outcome measures

We used the method proposed by Nanditha et al^21^ to obtain estimates of prevalence. For any IC, if a person has (at least) one visit with the IC during the year of analysis, then the lookback period is considered as follows:

- If the person also has at least one visit with the same IC during the lookback period, then this case is defined as a **prevalent** case;
- (Conversely, if the person does not have a visit with the same IC during the lookback period, then this case is defined as an **incident** case).

The lookback period is a period defined retrospectively as ending in the year of analysis and starting in the year equal to year of analysis minus L, where L, the value of lookback period, is chosen to be equal to 14. The year of analysis was 2022. Therefore, the lookback period started in 2008.

From the results produced using the DAD, one can apply assumptions on the *hospitalization rates* to deduce the general population prevalence rate of ICs. The reasoning is the following: if, for a given IC, T is the (unobserved) general population prevalence rate and H is the hospitalization rate, then the observed rate will be R=T*H. From this, one deduces that T=R/H. We used different values of H (varying from 0.1 to 1) to obtain estimates of T.

## III Outcomes

### 1. Global results

Using all diagnoses, we observe an overall prevalence rate of 0.18% among hospitalized individuals in Canada (0.18% for females and 0.19% for males) – see Table 2. If we make a conservative assumption that the global hospitalization rate is 0.1 (for all ICs), the estimates of the general population prevalence rate of ICs in Canada is about 1.8% (1.8% for females and 1.9% for males), which is comparable to the prevalence rates observed elsewhere^6,7,8^. Evidence-based hospitalization rates for specific ICs should be used in future investigations in order to obtain more precise prevalence estimates. We also observe that prevalence increases generally with age (except for PID).

**Table 2:**
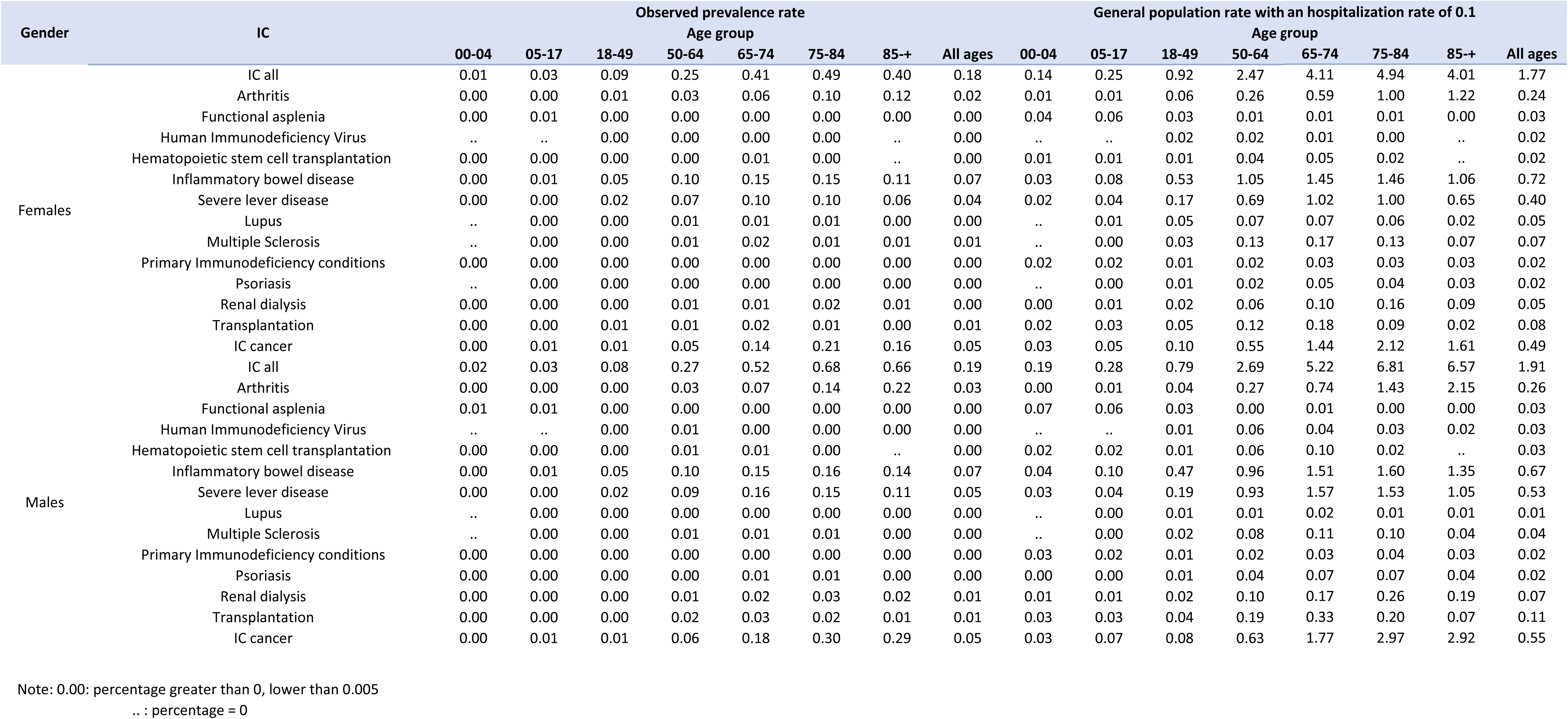
Prevalence of Immunodeficient conditions (in percentages) by definition, gender, age group: observed and general with hospitalization rate of 0.1, lookback period of 14 years.

| Gender | IC | Observed prevalence rate |  |  |  |  |  |  | General population rate with |  |  |  |  |
| --- | --- | --- | --- | --- | --- | --- | --- | --- | --- | --- | --- | --- | --- |
|  |  | Age group |  |  |  |  |  |  | Age g |  |  |  |  |
|  |  | 00-04 | 05-17 | 18-49 | 50-64 | 65-74 | 75-84 | 85-+ | All ages | 00-04 | 05-17 | 18-49 | 50-64 |
| Females | IC all | 0.01 | 0.03 | 0.09 | 0.25 | 0.41 | 0.49 | 0.40 | 0.18 | 0.14 | 0.25 | 0.92 | 2.47 |
|  | Arthritis | 0.00 | 0.00 | 0.01 | 0.03 | 0.06 | 0.10 | 0.12 | 0.02 | 0.01 | 0.01 | 0.06 | 0.26 |
|  | Functional asplenia | 0.00 | 0.01 | 0.00 | 0.00 | 0.00 | 0.00 | 0.00 | 0.00 | 0.04 | 0.06 | 0.03 | 0.01 |
|  | Human Immunodeficiency Virus | .. | .. | 0.00 | 0.00 | 0.00 | 0.00 | .. | 0.00 | .. | .. | 0.02 | 0.02 |
|  | Hematopoietic stem cell transplantation | 0.00 | 0.00 | 0.00 | 0.00 | 0.01 | 0.00 | .. | 0.00 | 0.01 | 0.01 | 0.01 | 0.04 |
|  | Inflammatory bowel disease | 0.00 | 0.01 | 0.05 | 0.10 | 0.15 | 0.15 | 0.11 | 0.07 | 0.03 | 0.08 | 0.53 | 1.05 |
|  | Severe lever disease | 0.00 | 0.00 | 0.02 | 0.07 | 0.10 | 0.10 | 0.06 | 0.04 | 0.02 | 0.04 | 0.17 | 0.69 |
|  | Lupus | .. | 0.00 | 0.00 | 0.01 | 0.01 | 0.01 | 0.00 | 0.00 | .. | 0.01 | 0.05 | 0.07 |
|  | Multiple Sclerosis | .. | 0.00 | 0.00 | 0.01 | 0.02 | 0.01 | 0.01 | 0.01 | .. | 0.00 | 0.03 | 0.13 |
|  | Primary Immunodeficiency conditions | 0.00 | 0.00 | 0.00 | 0.00 | 0.00 | 0.00 | 0.00 | 0.00 | 0.02 | 0.02 | 0.01 | 0.02 |
|  | Psoriasis | .. | 0.00 | 0.00 | 0.00 | 0.00 | 0.00 | 0.00 | 0.00 | .. | 0.00 | 0.01 | 0.02 |
|  | Renal dialysis | 0.00 | 0.00 | 0.00 | 0.01 | 0.01 | 0.02 | 0.01 | 0.00 | 0.00 | 0.01 | 0.02 | 0.06 |
|  | Transplantation | 0.00 | 0.00 | 0.01 | 0.01 | 0.02 | 0.01 | 0.00 | 0.01 | 0.02 | 0.03 | 0.05 | 0.12 |
|  | IC cancer | 0.00 | 0.01 | 0.01 | 0.05 | 0.14 | 0.21 | 0.16 | 0.05 | 0.03 | 0.05 | 0.10 | 0.55 |
| Males | IC all | 0.02 | 0.03 | 0.08 | 0.27 | 0.52 | 0.68 | 0.66 | 0.19 | 0.19 | 0.28 | 0.79 | 2.69 |
|  | Arthritis | 0.00 | 0.00 | 0.00 | 0.03 | 0.07 | 0.14 | 0.22 | 0.03 | 0.00 | 0.01 | 0.04 | 0.27 |
|  | Functional asplenia | 0.01 | 0.01 | 0.00 | 0.00 | 0.00 | 0.00 | 0.00 | 0.00 | 0.07 | 0.06 | 0.03 | 0.00 |
|  | Human Immunodeficiency Virus | .. | .. | 0.00 | 0.01 | 0.00 | 0.00 | 0.00 | 0.00 | .. | .. | 0.01 | 0.06 |
|  | Hematopoietic stem cell transplantation | 0.00 | 0.00 | 0.00 | 0.01 | 0.01 | 0.00 | .. | 0.00 | 0.02 | 0.02 | 0.01 | 0.06 |
|  | Inflammatory bowel disease | 0.00 | 0.01 | 0.05 | 0.10 | 0.15 | 0.16 | 0.14 | 0.07 | 0.04 | 0.10 | 0.47 | 0.96 |
|  | Severe lever disease | 0.00 | 0.00 | 0.02 | 0.09 | 0.16 | 0.15 | 0.11 | 0.05 | 0.03 | 0.04 | 0.19 | 0.93 |
|  | Lupus | .. | 0.00 | 0.00 | 0.00 | 0.00 | 0.00 | 0.00 | 0.00 | .. | 0.00 | 0.01 | 0.01 |
|  | Multiple Sclerosis | .. | 0.00 | 0.00 | 0.01 | 0.01 | 0.01 | 0.00 | 0.00 | .. | 0.00 | 0.02 | 0.08 |
|  | Primary Immunodeficiency conditions | 0.00 | 0.00 | 0.00 | 0.00 | 0.00 | 0.00 | 0.00 | 0.00 | 0.03 | 0.02 | 0.01 | 0.02 |
|  | Psoriasis | 0.00 | 0.00 | 0.00 | 0.00 | 0.01 | 0.01 | 0.00 | 0.00 | 0.00 | 0.00 | 0.01 | 0.04 |
|  | Renal dialysis | 0.00 | 0.00 | 0.00 | 0.01 | 0.02 | 0.03 | 0.02 | 0.01 | 0.01 | 0.01 | 0.02 | 0.10 |
|  | Transplantation | 0.00 | 0.00 | 0.00 | 0.02 | 0.03 | 0.02 | 0.01 | 0.01 | 0.03 | 0.03 | 0.04 | 0.19 |
|  | IC cancer | 0.00 | 0.01 | 0.01 | 0.06 | 0.18 | 0.30 | 0.29 | 0.05 | 0.03 | 0.07 | 0.08 | 0.63 |

Note: 0.00: percentage greater than 0, lower than 0.005 .. : percentage = 0

| Age group | Annual hospitalization rate of 0.1 |  |  |  |
| --- | --- | --- | --- | --- |
|  | 65-74 | 75-84 | 85-+ | All ages |
|  | 4.11 | 4.94 | 4.01 | 1.77 |
|  | 0.59 | 1.00 | 1.22 | 0.24 |
|  | 0.01 | 0.01 | 0.00 | 0.03 |
|  | 0.01 | 0.00 | .. | 0.02 |
|  | 0.05 | 0.02 | .. | 0.02 |
|  | 1.45 | 1.46 | 1.06 | 0.72 |
|  | 1.02 | 1.00 | 0.65 | 0.40 |
|  | 0.07 | 0.06 | 0.02 | 0.05 |
|  | 0.17 | 0.13 | 0.07 | 0.07 |
|  | 0.03 | 0.03 | 0.03 | 0.02 |
|  | 0.05 | 0.04 | 0.03 | 0.02 |
|  | 0.10 | 0.16 | 0.09 | 0.05 |
|  | 0.18 | 0.09 | 0.02 | 0.08 |
|  | 1.44 | 2.12 | 1.61 | 0.49 |
|  | 5.22 | 6.81 | 6.57 | 1.91 |
|  | 0.74 | 1.43 | 2.15 | 0.26 |
|  | 0.01 | 0.00 | 0.00 | 0.03 |
|  | 0.04 | 0.03 | 0.02 | 0.03 |
|  | 0.10 | 0.02 | .. | 0.03 |
|  | 1.51 | 1.60 | 1.35 | 0.67 |
|  | 1.57 | 1.53 | 1.05 | 0.53 |
|  | 0.02 | 0.01 | 0.01 | 0.01 |
|  | 0.11 | 0.10 | 0.04 | 0.04 |
|  | 0.03 | 0.04 | 0.03 | 0.02 |
|  | 0.07 | 0.07 | 0.04 | 0.02 |
|  | 0.17 | 0.26 | 0.19 | 0.07 |
|  | 0.33 | 0.20 | 0.07 | 0.11 |
|  | 1.77 | 2.97 | 2.92 | 0.55 |

### 2. Specific results

To help focus the interpretation on important prevalence results, Table 3 presents the 5 most frequent ICs per gender and age groups. For all genders and all age groups, severe cancers, inflammatory bowel disease (IBD) and liver disease are among the most frequent ICs. PID is among the most frequent only in the youngest age group. Functional asplenia is among the most frequent in the two youngest age groups. In all groups except the youngest males, the youngest females and females aged 50 to 64, transplantation or renal dialysis is among the most frequent ICs. For females aged 50 to 64, multiple sclerosis is among the most frequent.

**Table 3:**
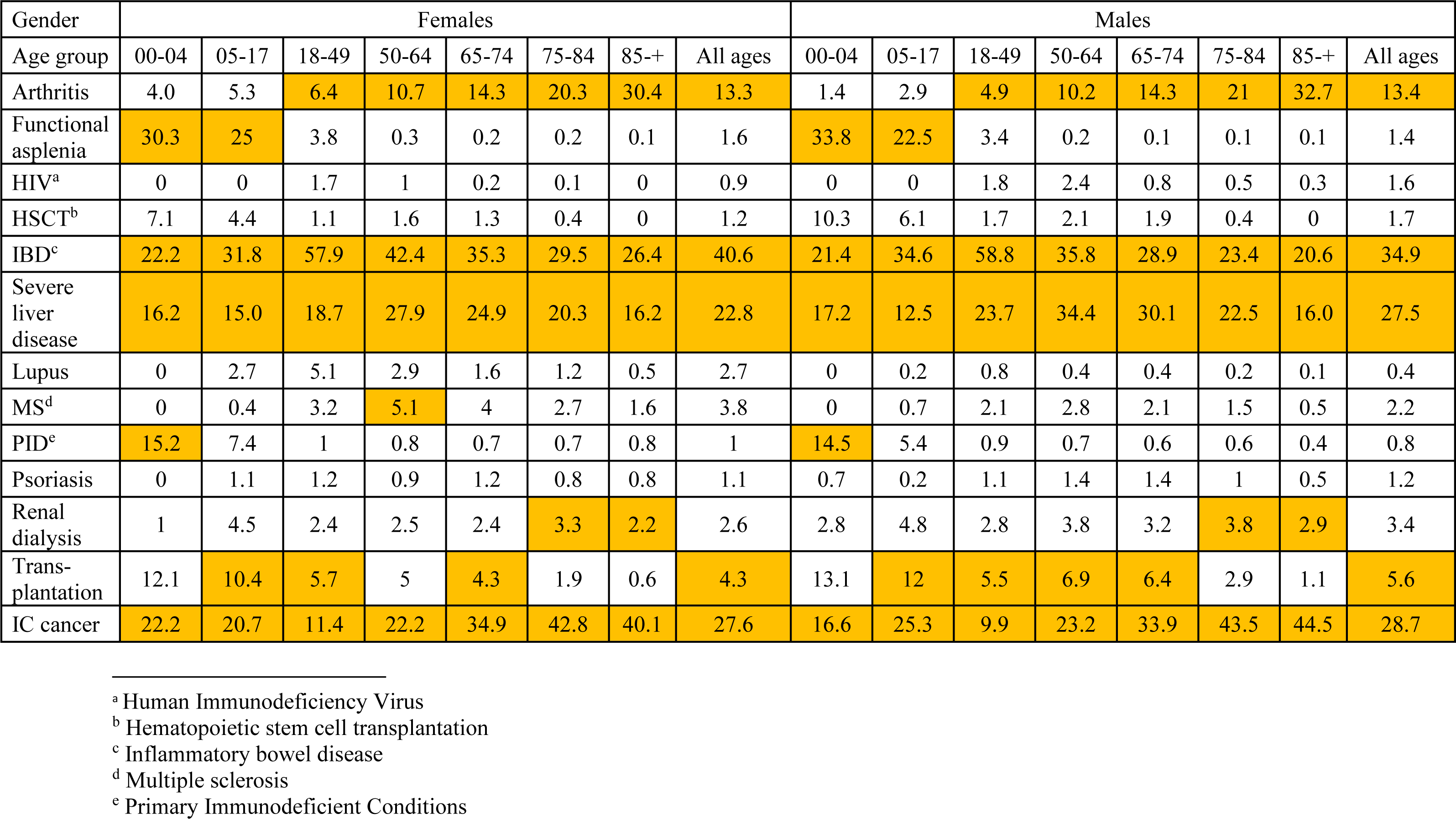
Percentage of specific ICs compared to total, by gender and age group, prevalence based on all diagnoses, lookback of 14 years, with yellow color indicating the 5 most frequent ICs per gender and age group.

## IV Discussion

### 1. Summary of key findings

ICs have an overall observed prevalence rate of about 0.18% at all in Canada, which corresponds to a total prevalence rate of about 1.8% if the hospitalization rate is uniformly 0.1.

### 2. Strengths and limitations

The DAD is limited to hospital visits. Any medical visit not performed in a hospital is not captured. We obtained prevalence rates that are therefore likely underestimates, but we tried to account for this by including a hospitalization rate (in the Excel file). For now, this rate has values 0.9, 0.75, 0.5, 0.25, 0.1, but, if needed, it will be straightforward for the user to include other values of hospitalization rates, for each IC, for each gender, and for each age group: a thorough literature review may help in this choice.

The DAD has been validated over the years. Nevertheless, it may be subject to misclassification^22^. Also, as mentioned earlier, it was not possible with the ICD-10 codes to consider all the nuances (e.g. simultaneous use of medications, transient nature of the IC, etc.) needed for the definition of an IC.

Definition of severe cancer proved challenging: it may differ depending on expert opinion or source. For this analysis, we performed a rudimentary search of the Canadian Cancer Society website to retrieve all the documentation that contained the word “Immunodeficiency”. The documents extracted mentioned the diseases that may be indicators of immunosuppression, according to at least one of the following criteria: HIV/AIDS, previous chemotherapy, previous radiation therapy, graft-versus-host disease after HSCT, weakened immune system (at birth or caused by immunosuppressant drugs).

Data from the province of Québec are not available in DAD. However, if needed, results may be generalized to Canada including Québec if it is assumed that similar trends to the rest of the country are observed in this province. Finally, the COVID-19 pandemic may have had an impact on hospitalization rates and consequently on the reliability of recent hospitalization data^23^.

### 3. Implications and next steps

Results may allow the user to determine the prevalence rates, given a priori estimates of hospitalization rates. Alternately, this project may be refined using hospitalization rates drawn from a more extensive review of literature to help obtain more accurate prevalence rates.

### 4. Conclusion

The general population prevalence rate (obtained when a hospitalization rate is conservatively assumed to be 0.1 uniformly) is about 1.8% overall, which is an indication on the potential vaccine recipients that may have an IC. The rate increases by age group in general and does not vary much between genders. The question remains on what should be the appropriate values of hospitalization rates specific to individual ICs, to obtain a more precise estimate of their global prevalence in Canada.

## Data Availability

All data produced in the present study are available upon reasonable request to the author

## Acknowledgments

The author thanks Elissa Abrams, Robert Pless, Marina Salvadori, Winnie Siu, and Linlu Zhao from PHAC for their advice in designing the study.

